# Genome-wide genetic overlap between fear-based disorders and generalised anxiety disorder

**DOI:** 10.64898/2026.02.06.26345742

**Authors:** Abigail R. ter Kuile, Brittany L. Mitchell, Sang Hyuck Lee, Geneviève Morneau-Vaillancourt, Megan Skelton, Jonathan R. I. Coleman, Helena L. Davies, Jessica Mundy, Alicia J. Peel, Christopher Hübel, Molly R. Davies, Anna E. Fürtjes, Zain Ahmad, Yuhao Lin, Brett N. Adey, Thomas McGregor, Alish Palmos, Johan Zvrskovec, Matthew Hotopf, Gursharan Kalsi, Ian R. Jones, Daniel J. Smith, David Veale, James T. R. Walters, Chérie Armour, Colette R. Hirsch, Andrew M. McIntosh, Naomi R. Wray, Sarah E. Medland, Enda M. Byrne, Nicholas G. Martin, Nathalie Kingston, John R. Bradley, NIHR BioResource, Gerome Breen, Thalia C. Eley

## Abstract

Twin studies reveal high genetic overlap between anxiety disorders and depression, contributing to the internalising spectrum. Some genetic specificity for fear-based anxiety disorders (fear), distinct from general anxiety and depression (distress), has also emerged. Limited datasets with detailed phenotyping across anxiety disorders have restricted most genome-wide association studies (GWAS) to “any anxiety diagnosis”. Additional genome-wide evidence to discern genetic differences between fear and distress is required. We conducted GWAS meta-analyses of fear (panic, agoraphobia, specific phobia, social anxiety disorder) and generalised anxiety disorder (GAD), measured using brief single-item and detailed symptom-based diagnoses from three datasets. We explored two control group criteria: phenotype-specific (fear/GAD) or broader anxiety/depression screening. We identified one SNP-based independent locus and three gene-level genome-wide significant (GWS) associations with fear (up to 35,523 N_cases_; 157,447 N_controls_). Four GWS SNP-based loci and three gene-level loci were associated with GAD (up to 60,879 N_cases_; 117,064 N_controls_). The genetic correlation between fear and GAD was significantly different from unity only when excluding a depression-enriched dataset and using phenotype-specific control screening (r_g_ = 0.87; P = 9.32 × 10^−3^). Most complex traits had statistically similar genetic correlations with fear and GAD, including depression. Exceptions included general cognitive ability, educational attainment, and coronary artery disease, showing statistically stronger genetic correlations with fear than GAD, while bipolar disorder type I, anorexia nervosa, and neuroticism displayed the opposite pattern. Our findings partially support a distress-fear genetic distinction, but show stronger evidence for an overarching genetic liability to internalising psychopathology driving comorbidity across anxiety disorders and depression.

## Introduction

While high comorbidity exists across anxiety and depressive disorders [1–3], their distinct features form the basis of a model separating fear and distress [4,5]. Fear-based anxiety disorders (panic disorder, agoraphobia, specific phobia, and social anxiety disorder) are characterised by context-specific fearful arousal and behavioural avoidance of a narrow range of stimuli [4,5]. In contrast, generalised anxiety disorder (GAD) presents as uncontrollable worry about a broader range of situations. GAD is therefore often categorised alongside depression as a distress-based disorder, sharing core features of broad, pervasive negative emotions and exhibiting higher comorbidity with each other than with fear-based disorders [4–6]. Prior studies suggest an overarching shared liability alongside subtype-specific risk factors across distress-fear dimensions [5].

Twin studies estimate anxiety heritability in the range 20-60% [7–9], with evidence of genetic influences contributing to a broad internalising liability shared among individual anxiety disorders and depression [10–13]. However, other findings suggest this higher-order genetic liability can be differentiated into replicable lower-order genetic components. Fear-based disorders show partial genetic distinction from GAD, as a distress-based disorder, resulting in two distinct but correlated subfactors underlying anxiety genetics [10,14].

While twin studies can examine the genetic architecture of the anxiety disorder spectrum, molecular genetic approaches are needed to identify the specific genetic variants involved. Genome-wide association studies (GWAS) have begun identifying common genetic variants underpinning anxiety disorders, though fewer anxiety studies exist compared to other psychiatric disorders [15]. Recent large-scale GWAS of broadly-defined anxiety and GAD symptoms [16–20], and one smaller panic disorder GWAS [21], revealed significant genetic correlations between anxiety and other psychiatric conditions, including depression, schizophrenia, bipolar disorder, and attention deficit hyperactivity disorder (ADHD) [16,18–21]. Consistent with twin studies [10], preliminary genome-wide findings suggested that self-reported single-item diagnoses of any fear-based disorder (as a group) showed weaker genetic correlations with depression than between depression and GAD [22]. However, the common genetic variants associated with fear-based disorders remain underexplored, limiting our molecular-level understanding of the internalising genetic spectrum.

A previous panic disorder GWAS (∼2,250 cases) found a substantial genetic correlation with a combined any anxiety disorder phenotype, although this did not survive multiple testing correction [21]. This and other fear-based disorder GWAS were underpowered for loci discovery or assessment of genetic correlations with a sufficiently broad range of traits to understand where fear disorders sit genetically in relation to other traits [22–24]. Formal comparisons of fear and GAD genetic correlations with other psychiatric, behavioural, cognitive, and health traits could help identify transdiagnostic or subtype-specific mechanisms. Such analyses would improve molecular genetic evidence and fine-grained detail of psychiatric spectrum hierarchical structures, with the potential to improve diagnostic systems [5]. Further well-powered genome-wide evidence of the distress-fear genetic distinction is needed in the broader context of their relationships with other complex traits.

Anxiety disorder phenotyping is an important consideration when identifying genetic similarities and differences. Addressing comorbidity is a particular challenge in identifying anxiety-specific genetic influences, as anxiety disorders are among the most highly comorbid disorders across psychopathology [25]. Including heritable comorbid traits as GWAS covariates can bias results [26]. Screening controls for co-occurring traits in GWAS could inflate genetic correlations [27], necessitating the comparison of different screening approaches. Furthermore, genetic influences captured using brief phenotyping may be less specific to the focal trait than those from detailed phenotyping, reflecting more general psychopathology [28,29]. Previous GWAS reporting distress-fear genetic correlations relied on brief, single-item reports of a diagnosis by a health professional for each fear disorder [22]. We previously showed that lower rates of fear-based disorder diagnoses were reported via brief self-reports than when using detailed symptom-based measures, with the opposite pattern for GAD [30]. Identifying genetic variants requires large sample sizes, creating a trade-off between maximising sample size and phenotypic detail. Combining detailed symptom-based diagnoses with brief single-item self-report diagnoses can achieve larger sample sizes while retaining phenotypic specificity [31].

In this study, we assessed shared and non-shared genetic influences on fear-based disorders and GAD at the level of common genetic variants. We increased sample size and power for loci-discovery and genetic correlation analyses by combining detailed symptom-based diagnoses with brief single-item diagnoses. We examined differences in genetic correlations between fear-based disorders and GAD with a range of other complex traits and assessed broad and specific approaches to screening controls. Our study represents significant progress in anxiety disorder genomics and provides further detail on the distress-fear distinction at the genome-wide level.

## Methods

### Datasets and measures

#### Overview

We conducted GWAS meta-analyses of two lifetime anxiety disorder phenotypes i) fear-based disorders (at least one of panic, agoraphobia, specific phobia or social anxiety disorder; ‘fear’; N cases = 35,523) and ii) GAD (N cases = 60,879). We combined detailed symptom-based diagnoses and brief, self-report diagnoses from six samples, resulting in three case-control datasets (**Table 1 & Supplementary Table 1**).

**Table 1.** Sample sizes in each GWAS dataset and meta-analysis of fear-based disorders and GAD.

| Anxiety disorder | GWAS Datasets | N<sub>cases</sub> | Control criteria |  |  |  | N dataset total<br>(N<sub>cases</sub> + N<sub>controls</sub>) |  |
| --- | --- | --- | --- | --- | --- | --- | --- | --- |
|  |  |  | Anx-dep controls |  | Specific controls |  | Anx-dep | Specific |
|  |  |  | N<sub>controls</sub> | N<sub>effective</sub> | N<sub>controls</sub> | N<sub>effective</sub> |  |  |
| Fear | GLAD+ | 18,091 | 4,002 | 13,108 | 5,693 | 17,321 | 22,093 | 23,784 |
|  | UKB | 10,789 | 83,236 | 38,204 | 137,209 | 40,010 | 94,025 | 147,998 |
|  | QIMR | 6,643 | 12,722 | 17,457 | 14,545 | 18,241 | 19,365 | 21,188 |
|  | 'Restricted' meta-analysis (GLAD+ & UKB) | 28,880 | 87,238 | 51,312 | 142,902 | 57,331 | **116,118** | **171,782** |
|  | 'Full' meta-analysis (GLAD+, UKB & QIMR) | 35,523 | 99,960 | 68,769 | 157,447 | 75,572 | **135,483** | **192,970** |
| GAD | GLAD+ | 21,984 | 4,002 | 13,543 | 6,804 | 20,784 | 25,986 | 28,788 |
|  | UKB | 26,067 | 83,236 | 79,402 | 95,715 | 81,950 | 109,303 | 121,782 |
|  | QIMR | 12,828 | 12,722 | 25,550 | 14,545 | 27,265 | 25,550 | 27,373 |
|  | 'Restricted' meta-analysis (GLAD+ & UKB) | 48,051 | 87,238 | 92,944 | 102,519 | 102,733 | **135,289** | **150,570** |
|  | 'Full' meta-analysis (GLAD+, UKB & QIMR) | 60,879 | 99,960 | 118,494 | 117,064 | 129,999 | **160,839** | **177,943** |

Numbers shown in bold are total sample size used in GWAS meta-analyses. Effective sample size was calculated as: $N_{\text{effective}} = 4 / [(1/n_{\text{case}}) + (1/n_{\text{control}})]$ , which converts the power of a study sample size to one with a balanced case-control ratio (i.e. equivalent to a study with a sample prevalence of 50%, with $N_{\text{cases}} = N_{\text{effective}}$ and $N_{\text{controls}} = N_{\text{effective}}$ ).

#### GLAD+

The GLAD+ dataset included three samples, where cases were primarily from the Genetic Links to Anxiety and Depression (GLAD) Study, which recruited UK participants with a lifetime anxiety and/or depressive disorder [32]. Additional cases were from the United Kingdom Eating Disorders Genetics Initiative (EDGI UK) [33] and the COVID-19 Psychiatric and Neurological Genetics (COPING) Study (**Supplementary Materials**) [34,35]. The GLAD+ dataset used brief and detailed measures of all five anxiety disorders to define cases and controls. Controls were ascertained from the COPING Study [34,35].

#### QIMR Berghofer

The Queensland Institute of Medical Research (QIMR) Berghofer dataset comprised cases from the Australian Genetics of Depression Study (AGDS) [36] and controls from the population-based QIMR Berghofer QSkin Sun & Health Study [37]. AGDS used detailed and brief measures, while QSkin used brief measures only. As all recruited AGDS participants had lifetime depression, this might increase depression/distress genetics in anxiety disorder GWAS, thus elevating the genetic correlation between fear, GAD, and depression. To address this potential depression-enrichment bias in anxiety cases, we conducted secondary ‘*restricted*’ meta-analyses excluding QIMR.

#### UK Biobank

The UK Biobank (UKB) is a population health cohort study [38] from which our dataset included a subset of participants who completed the Mental Health Questionnaire [39] and provided brief measures for fear disorders and both brief and detailed measures for GAD. We excluded GLAD+ participants who were also in UKB to prevent sample overlap.

#### Case-control definitions

We derived detailed symptom-based anxiety disorder measures using previously described algorithms [30]. Cases met DSM-5 criteria for a lifetime symptom-based disorder diagnosis, assessed using an online, self-report version of the Composite International Diagnostic Interview short-form (CIDI-SF) [36,40]. Our brief measures comprised single-item self-report questions (e.g. “Have you ever been diagnosed with one or more of the following mental health problems by a professional?”; **Supplementary Table 2**). For panic assessments, we used the broad term “panic” to describe case-ness as we included both detailed diagnostic measures of panic disorder and brief measures of panic attacks. Although panic attacks are not specific to panic disorder, the brief diagnostic measure of panic attacks has shown agreement with detailed diagnostic measures of panic disorder [30].

We explored two control group definitions. First, controls screened for any anxiety disorder or depression (fear_anx-dep_ and GAD_anx-dep_). Due to the high genetic correlation between these disorders [22], this was the most powerful approach for loci discovery. Second, specifically screened controls (fear_specific_ and GAD_specific_) whereby participants were only screened for the specific disorder being analysed to better distinguish genetic specificity between GAD and fear. Specific screening of controls was not possible in the QIMR dataset as we were limited to using a broad, single-item self-report diagnosis of “anxiety” (**Supplementary Tables 1** & **2**). Controls were not screened for the presence of other psychiatric disorders as this can bias genetic correlation estimates [27].

In summary, we conducted four GWAS meta-analyses with two control definitions (fear_anx-dep_, GAD_anx-dep_, fear_specific_ and GAD_specific_) using both the *‘full’* dataset (all three GWAS datasets) and the *‘restricted’* dataset (excluding the depression-enriched QIMR dataset). This resulted in eight meta-analyses in total.

### Genome-wide association analyses

Genotyping and quality control were performed separately in each study (**Supplementary Materials**). Common genetic variants were imputed to the TOPMed imputation panel Version R2 on GRC38 (GLAD+ and QIMR) or the Haplotype Reference Consortium and UK10K Consortium reference panels (UKB). Analyses were restricted to common variants (MAF > 1%) with imputation confidence INFO scores > 0.3 for TOPMed or > 0.4 for UKB.

We conducted four anxiety disorder GWASs: fear_specific_, GAD_specific_, fear_anx-dep_ and GAD_anx-dep_. Prior to meta-analysis, we performed a separate GWAS for these four phenotypes in each of our three datasets (GLAD+, QIMR, UKB; **Table 1**) using REGENIE v3.1.3 (GLAD+, UKB) or SAIGE v0.44 (QIMR) [41,42]. Covariates included the first ten principal components, genotyping batch, array (GLAD+ only), and assessment centre (UKB only). Analyses were limited to participants of European-associated genetic ancestry clusters, defined using principal component analysis (PCA). Prior to meta-analysis, GWAS summary statistics were processed and harmonised using the MungeSumstats R package (**Supplementary Materials**) [43].

### Meta-analyses and annotation

We used METAL to perform inverse-variance weighted meta-analyses of each anxiety disorder phenotype [44]. Primary meta-analyses (‘*full’)* included all three datasets, while secondary meta-analyses (‘*restricted’*) excluded QIMR to address potential depression-enrichment bias. We restricted common genetic variants to those overlapping across datasets in each meta-analysis. Meta-analysis results were mapped and annotated using FUMA v1.5.2 with default parameters applied and the “UKB release2b 10K European” reference panel [45]. MAGMA v1.08 with default parameters was used for gene-level and biological pathway analyses [46].

### SNP-based heritability

We estimated the liability-scale heritability captured by common genetic variants (h^2^) of fear and GAD using LDSC regression [47]. We used the sum of effective sample size to adjust for variability in sample prevalence across datasets [48]. Population prevalences were derived from the COPING study, in which both detailed and brief measures were available to define cases and controls for all five anxiety disorders (**Supplementary Table 3**). Although not population-based, COPING study prevalence estimates aligned with epidemiological ranges for individual anxiety disorders [49,50]. Given the lack of epidemiological studies estimating the population prevalence of fear-based disorders as a group, we also calculated h^2^_SNP_ liability-scale estimates using prevalences 5% higher and lower than the COPING study prevalences.

### Genetic correlations

Using LDSC regression [51], we calculated genetic correlations (r_g_) between i) fear and GAD and ii) fear and GAD with over a hundred external complex traits. Traits were considered sufficiently powered if they had an LDSC GWAS mean ꭓ^2^ > 1.02 and heritability Z score > 4 [52]. We tested if the fear-GAD genetic correlation was significantly different from 0 using default parameters in bivariate LDSC regression, and significantly different from 1, calculated in R using the chi-squared distribution function and [(|r_g_|−1)/se]^2^. To correct for multiple testing, we applied a Bonferroni corrected *P*-value threshold (*α*/number of fear-GAD r_g_ tested; 0.05/4 = *P* ≤ 0.0125). Fear and GAD GWAS with a genetic correlation significantly different from 1 were then tested for genetic correlations with external traits and applied a Bonferroni-corrected significance threshold (*α*/number of external traits tested; 0.05/106 = *P* ≤ 4.71 × 10^−4^). For external traits with significant genetic correlations with GAD or fear, we tested if these correlations were statistically different between fear versus GAD using the block-jackknife LDSC method [51,53]. Significant differences in genetic correlations between fear and GAD with external traits were determined using the P-value derived from the block-jackknife Z statistic, with a Bonferroni-corrected threshold adjustment for all external traits tested for r_g_ (*P* ≤ 4.71 × 10^−4^).

## Results

### Genome-wide association meta-analyses

Manhattan plots for the most well-powered GWAS meta-analysis are shown in **Figure 1** (fear_anx-dep_ and GAD_anx-dep_ in the primary *full* meta-analysis; see **Supplementary Figure 1** for Q-Q plots). Manhattan plots and Q-Q plots for all other meta-analyses (fear_specific_, GAD_specific_ and the secondary *restricted* meta-analyses) are shown in **Supplementary Figures 1-4**. We found little evidence of confounding in all GWAS meta-analyses (LDSC intercept = 1.00-1.01; **Supplementary Table 4**). LDSC intercepts >1 indicate some inflation, which is expected as the GWAS sample size increases [54]. The majority of inflation in our meta-analyses was due to polygenicity (93-100%), as indicated by LDSC attenuation ratio calculations (**Supplementary Table 4)**. Regional plots for independent genome-wide significant loci are shown in **Supplementary Figures 5-9**.

**Figure 1:**
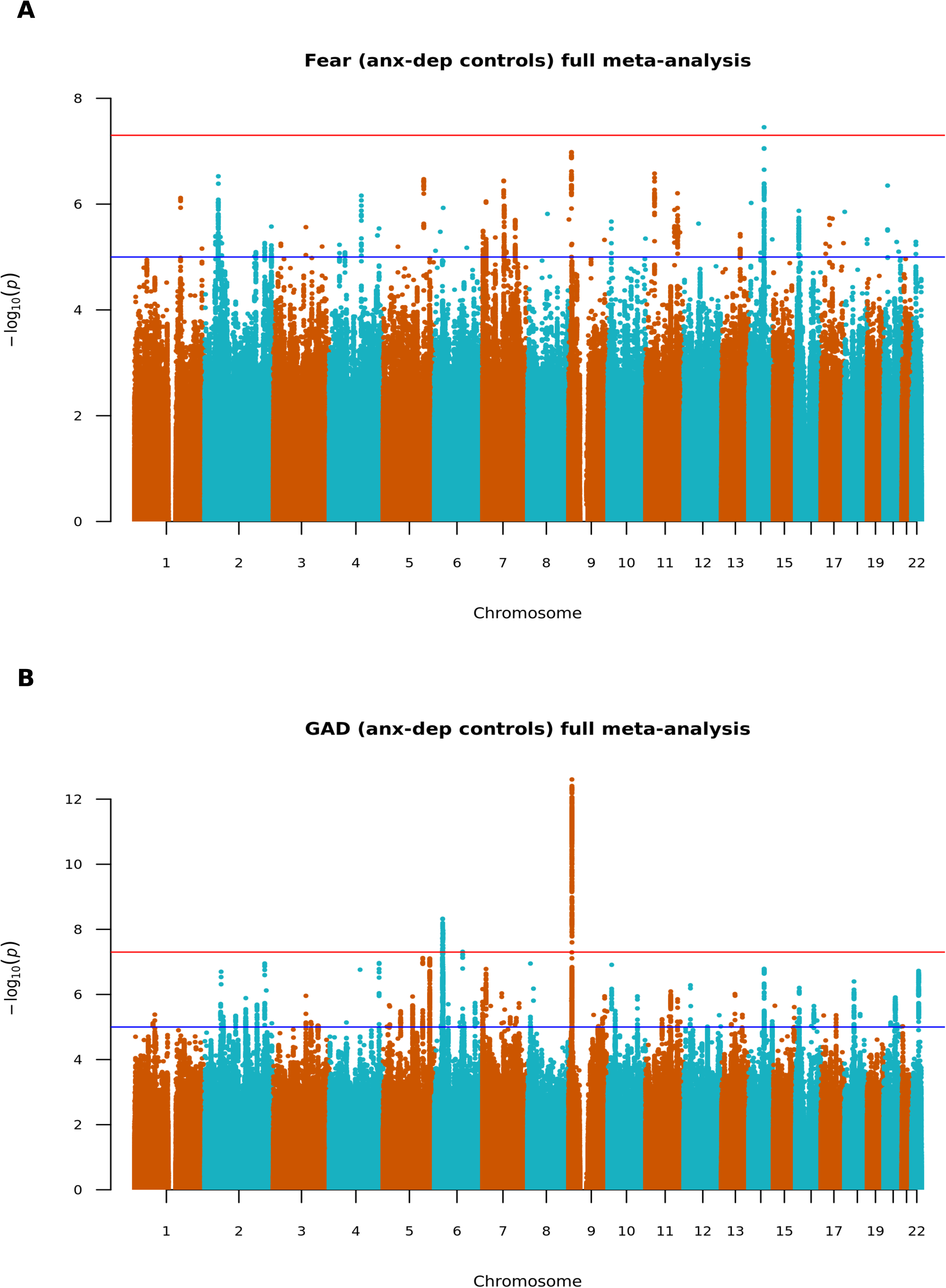
Genome-wide association study Manhattan plots for anxiety disorder phenotypes in the ‘*full*’ meta-analyses (GLAD+, UKB & QIMR datasets). Fear-based disorder GWAS (A) and generalised anxiety disorder GWAS (B) results from phenotypes with controls screened for any anxiety disorder and depression. Line red; common genetic variant genome-wide significance threshold (P ≤ 5 × 10^−8^), blue; suggestive significance threshold (P ≤ 1 × 10^−5^).

We identified one independent genome-wide significant locus (*P* ≤ 5 × 10^−8^) associated with fear in the *full* fear_anx-dep_ meta-analysis (lead variant rs10047892 on chromosome 14; *P* = 3.52 × 10^−8^; **Figure 1A**; **Table 2**). No loci reached *P* ≤ 5 × 10^−8^ in the three other fear meta-analyses (fear_specific_ in *full*, or fear_specific_ and fear_anx-dep_ in *restricted*). Across our GAD meta-analyses, we identified four independent genome-wide significant loci associated with GAD. The most significant was on chromosome 9 in the GAD_anx-dep_ *full* meta-analysis (9p23; lead variant rs17189482; *P* = 2.49 × 10^−13^; **Figure 1B**) and was genome-wide significant in all four GAD meta-analyses (**Table 2**). The locus on chromosome 6 (6p22.1) was the second most significant in GAD_anx-dep_ *full* meta-analysis (lead variant rs385816; *P* = 4.73 × 10^−9^), followed by locus 6q16.3 (lead variant rs17185536; *P* = 4.88 × 10^−8^; **Figure 1, lower panel).** An additional locus on chromosome 2 (2p16.1) exceeded genome-wide significance in GAD_specific_ *full* meta-analysis (lead variant rs11688767; *P* = 3.53 × 10^−8^; **Table 2**).

**Table 2:**
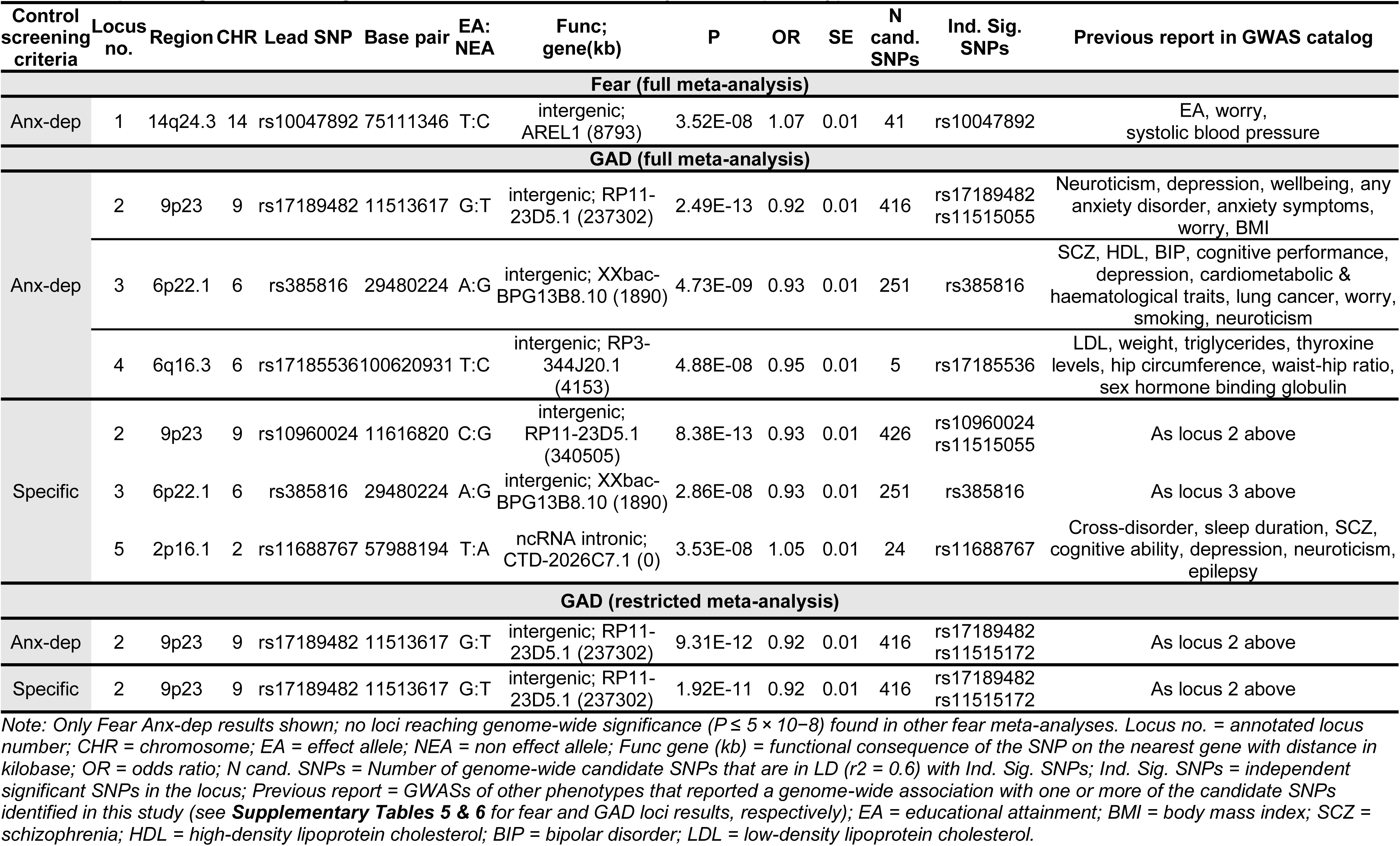
Independent genome-wide significant loci associated with anxiety disorder phenotypes.

### Gene-level and gene-set association analyses

Three genes and two gene-sets were significantly associated with fear. *SNX29* (chromosome 16) was significantly associated with fear in all four meta-analyses. *PAX6* (chromosome 11) was significant in fear_anx-dep_ *full* meta-analysis, while *ERI3* (chromosome 1, includes the SNP-level genome-wide significant locus 1p34.1) was significant in fear_anx-dep_ *restricted* meta-analysis (**Table 3**). The gene-set suppression of apoptosis [*P* = 7.89 × 10^−10^; Bonferroni correction threshold for 17008 gene-sets = *P* ≤ 2.94 × 10^−6^] was associated with fear_specific_ and fear_anx-dep_ *full* meta-analyses. Negative regulation of cell growth involved in cardiac muscle development [*P* = 3.32 × 10^−7^] was associated with fear_specific_ *full* meta-analysis (**Supplementary Table 7**).

**Table 3:** Gene-level associations with anxiety disorder phenotypes.

| Control screening criteria | Gene symbol | CHR | N SNPS | Z | P | P Bonf. Threshold (0.05/N genes) | Previous report in GWAS catalog | Gene name |
| --- | --- | --- | --- | --- | --- | --- | --- | --- |
| **Fear (full meta-analysis)** |  |  |  |  |  |  |  |  |
| Anx-dep | SNX29 | 16 | 2615 | 5.3 | 5.66E-08 | 2.68E-06 (18664 genes) | EA, insomnia, smoking, cognitive ability, depression, HDL, type 2 diabetes | Sorting nexin 29 |
|  | PAX6 | 11 | 42 | 4.69 | 1.38E-06 |  | Neuroticism, depression, worry, blood pressure, BMI, insomnia, irritability, wellbeing | Paired box 6 |
| Specific | SNX29 | 16 | 2616 | 5.27 | 6.94E-08 |  | As above | As above |
| **Fear (restricted meta-analysis)** |  |  |  |  |  |  |  |  |
| Anx-dep | SNX29 | 16 | 2618 | 5.41 | 3.22E-08 | 2.68E-6 (18675 genes) | As above | As above |
|  | ERI3 | 1 | 192 | 4.69 | 1.34E-06 |  | EA, alcohol consumption, height | ERI1 exoribonuclease family member 3 |
| Specific | SNX29 | 16 | 2620 | 5.27 | 6.64E-08 |  | As above | As above |
| **GAD (full meta-analysis)** |  |  |  |  |  |  |  |  |
| Anx-dep | TMEM106B | 7 | 195 | 4.78 | 8.84E-07 | 2.68E-6 (18665 genes) | Anxiety, depression, EA, wellbeing, insomnia, Alzheimer's, neuroticism, GAD symptoms, PTSD, HDL | Transmembrane protein 106B |
|  | YLPM1 | 14 | 155 | 4.67 | 1.47E-06 |  | Anxiety, EA, neuroticism, blood pressure, HDL, body fat %, cortical thickness | YLP motif-containing protein 1 |
|  | SORCS3 | 10 | 1818 | 4.6 | 2.16E-06 |  | Anxiety, EA, depression, blood pressure, externalising behaviours, CAD, insomnia, neuroticism, smoking, ADHD | Sortilin related VPS10 domain containing receptor 3 |

Gene-level association analyses conducted in MAGMA. Only analyses exceeding Bonferroni-corrected significance thresholds shown. CHR = chromosome; Previous report = GWASs of other phenotypes that report genome-wide significant loci that map onto the gene-level associations identified in this study (see **Supplementary Tables 8 & 9** for fear and GAD results, respectively); EA = educational attainment; SCZ = schizophrenia; GAD = generalised anxiety disorder; PTSD = post-traumatic stress disorder; HDL = high-density lipoprotein cholesterol; CAD = coronary artery disease; ADHD = attention deficit hyperactivity disorder.

Three genes were significantly associated with GAD in GAD_anx-dep_ *full* meta-analysis (*TMEM106B* on chromosome 7; *YLPM1* on chromosome 14; *SORCS3* on chromosome 10; **Table 3**). The YLPM1 gene was on the same locus as the genome-wide significant SNP-level association with fear (14q24.3). No genes were genome-wide significant in the three other GAD meta-analyses. No gene-sets were significantly associated with GAD after Bonferroni adjustment for 17008 gene-sets tested (*P* ≤ 2.94 × 10^−6^).

### SNP-based heritability

We report results here for the most powerful GWAS (fear_anx-dep_ and GAD_anx-dep_ in the primary *full* meta-analyses), as measured by GWAS mean ꭓ^2^. We identified a liability-scale LDSC h^2^_SNP_ for fear_anx-dep_ of 12.9% (SE = 0.9%, mean ꭓ^2^ = 1.17) and 12.2% for GAD_anx-dep_ (SE = 0.6%, mean ꭓ^2^ = 1.25), assuming a population prevalence of 8.8% and 12.9%, respectively. **Supplementary Table 4** shows h^2^_SNP_ liability-scale estimates calculated from prevalences 5% higher and lower than COPING study sample prevalences. **Supplementary Table 4** also reports h^2^_SNP_ liability-scale estimates for phenotypes with specifically screened controls, and in the *restricted* meta-analysis dataset.

### Genetic correlation between fear-based disorders and GAD

The genetic correlation between fear and GAD was high (*r*_g_= 0.87-0.98) and significantly different from 0 for both phenotypes (fear_specific_–GAD_specific_ and fear_anx-dep_–GAD_anx-dep_) in both primary *full* and secondary *restricted* meta-analysis datasets. Given this high genetic overlap, we also tested whether these correlations significantly differed from unity. We found that only the genetic correlation between fear_specific_–GAD_specific_ in *restricted* meta-analyses was significantly less than 1 (i.e., controls were screened specifically and when the depression-enriched QIMR dataset was excluded from the meta-analysis). The other three fear–GAD genetic correlations were not significantly different from 1 (**Table 4**; Bonferroni-corrected significance threshold *P* ≤ 0.0125).

**Table 4:**
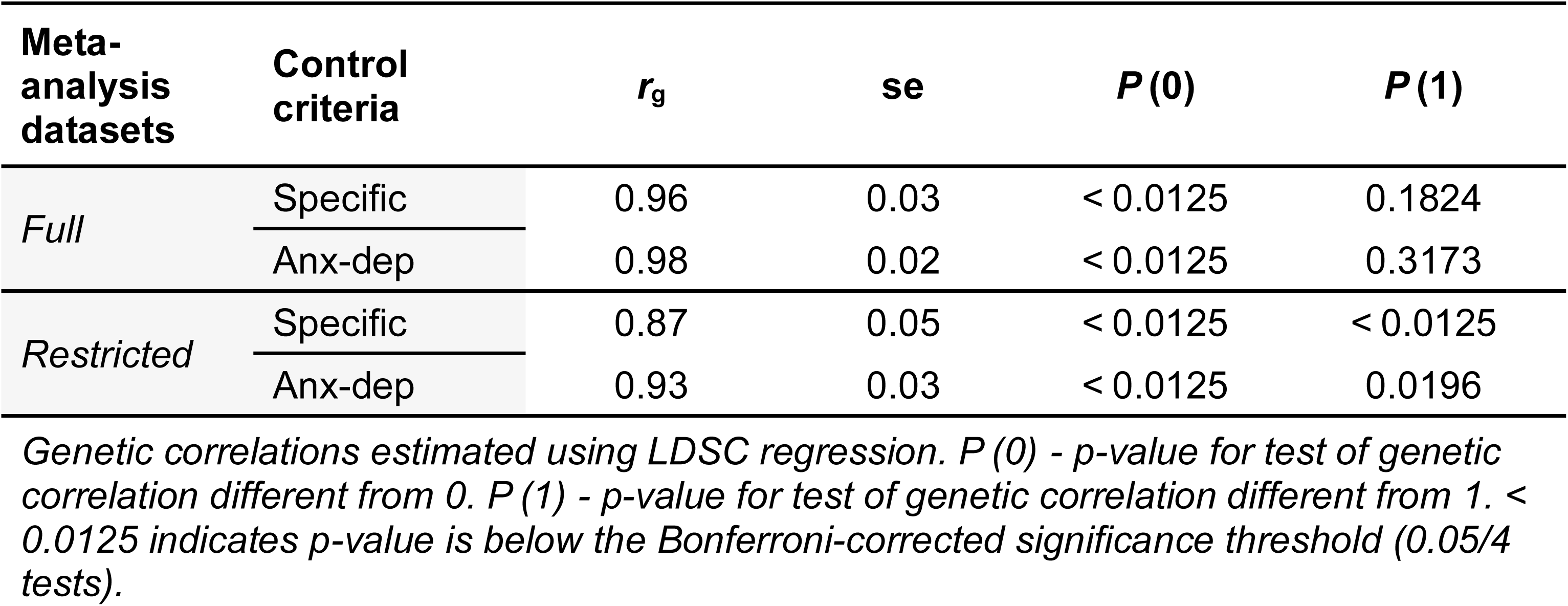
Genetic correlations (*r*_g_) between fear-based disorder and GAD phenotypes.

| Meta-analysis datasets | Control criteria | $r_g$ | se | $P(0)$ | $P(1)$ |
| --- | --- | --- | --- | --- | --- |
| *Full* | Specific | 0.96 | 0.03 | < 0.0125 | 0.1824 |
|  | Anx-dep | 0.98 | 0.02 | < 0.0125 | 0.3173 |
| *Restricted* | Specific | 0.87 | 0.05 | < 0.0125 | < 0.0125 |
|  | Anx-dep | 0.93 | 0.03 | < 0.0125 | 0.0196 |

*Genetic correlations estimated using LDSC regression. $P(0)$ - p-value for test of genetic correlation different from 0. $P(1)$ - p-value for test of genetic correlation different from 1. < 0.0125 indicates p-value is below the Bonferroni-corrected significance threshold (0.05/4 tests).*

### Genetic correlations with external phenotypes

We calculated genetic correlations between fear and GAD with over a hundred external phenotypes. Only fear_specific_ and GAD_specific_ in the *restricted* meta-analyses were tested for genetic correlations with external phenotypes (as it was only in this analysis that the fear–GAD genetic correlation was significantly different from 1). Most external traits showed similar patterns of genetic correlations with fear and GAD, where the genetic correlation between an external trait and fear_specific_ was not significantly different from its genetic correlation with GAD_specific_ (Bonferroni-corrected significance threshold *P* ≤ 4.71 × 10^−4^; **Figure 2; lower panel**). However, there were several notable exceptions. Educational attainment, general cognitive ability and cognitive aspects of educational attainment showed significantly stronger negative genetic correlations with fear_specific_ than with GAD_specific_. Coronary artery disease showed a significantly higher positive genetic correlation with fear_specific_ than with GAD_specific_. Conversely, a diagnosis of any bipolar disorder (type I and type II), bipolar disorder type I, anorexia nervosa, and neuroticism showed significantly stronger positive genetic correlations with GAD_specific_ than with fear_specific_ (**Figure 2; upper panel**). Full genetic correlation results with external phenotypes are shown in **Supplementary Table 10**.

**Figure 2:**
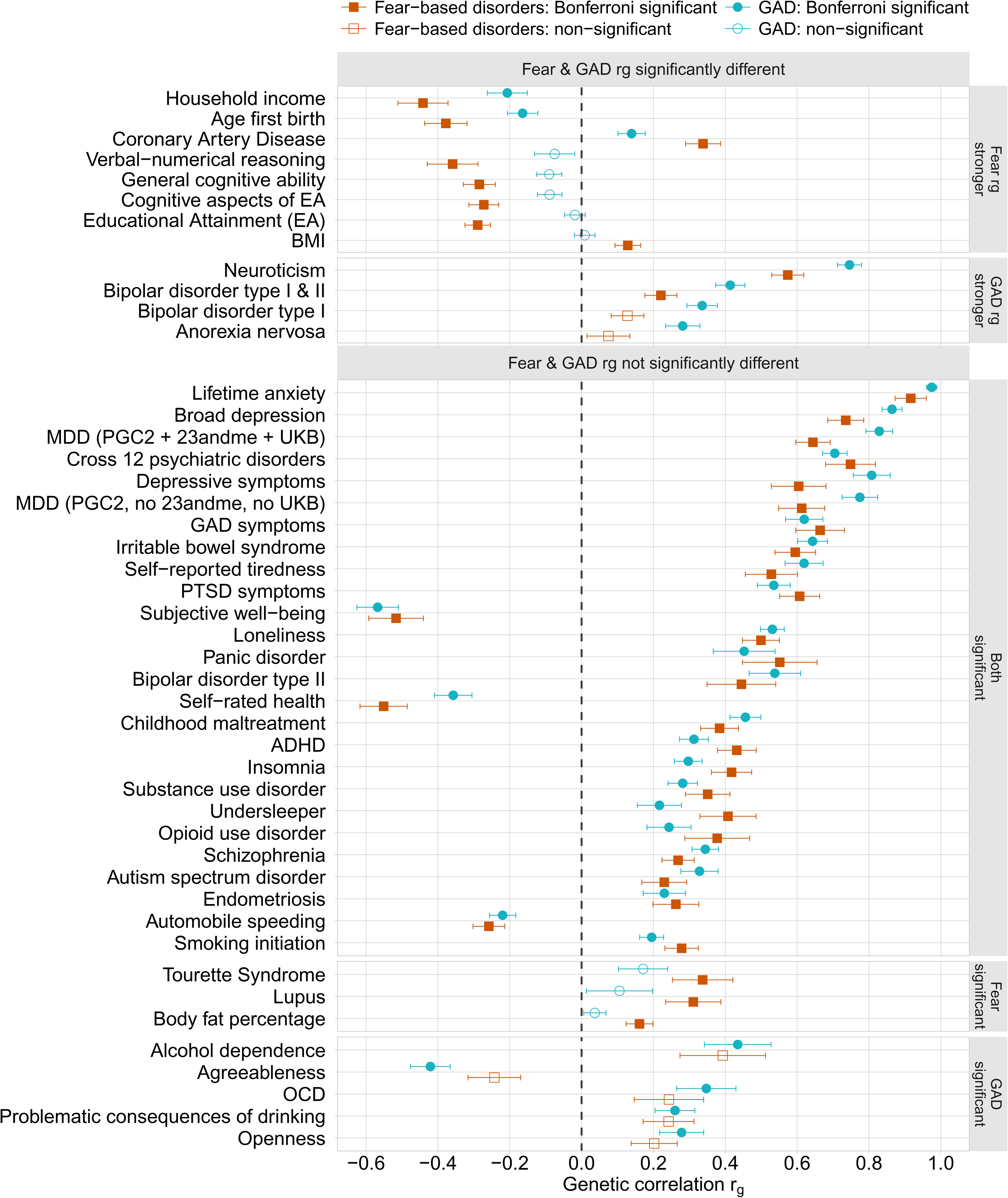
Genetic correlations between anxiety disorder phenotypes and external traits estimated in LDSC regression. Fear-based disorder GWAS and GAD GWAS results from restricted meta-analysis (GLAD+ and UKB datasets) with controls screened specifically for their respective anxiety disorders. Genetic correlations with 106 phenotypes were tested (Bonferroni-corrected significance threshold P ≤ 4.71 × 10^−4^). Differences between the genetic correlation of fear-based disorders and that of GAD with each external phenotype were tested using a block jackknife (Bonferroni correction for significance [P ≤ 4.71 × 10^−4^]). Only traits with genetic correlations significantly different from 0 with at least one of the anxiety disorder phenotypes are shown and were tested using block jackknife. Full results are shown in **Supplementary Table 10**. Upper panel: phenotypes with statistically significant different genetic correlations between fear-based disorders and GAD. Lower panel: phenotypes with no statistically significant different genetic correlations between fear-based disorders and GAD, though in the lower two segments, there was a significant association with one trait and not the other. Bars represent standard errors. ADHD = attention deficit hyperactivity disorder; BMI = body mass index; GAD = generalised anxiety disorder; MDD = major depressive disorder; OCD = obsessive-compulsive disorder; PGC = Psychiatric Genomics Consortium; PTSD = post-traumatic stress disorder; UKB = UK Biobank.

## Discussion

This study reports the first genome-wide significant (GWS) associations with any lifetime fear-based disorder diagnosis (fear). We identified one independent GWS SNP-based locus and three additional loci from gene-based analysis for fear, along with four GWS SNP-based loci and three genes associated with GAD. Combining detailed and brief diagnostic measures improved statistical power compared to previous fear GWASs [21,22]. Our study revealed high genetic correlations between fear, GAD, and depression, consistent with twin studies [10,12,13]. Excluding the depression-enriched dataset and only screening controls specifically for the anxiety disorder being analysed revealed some genetic specificity for fear and GAD. We identified significant differences in genetic correlations between fear and GAD with several traits, including cognitive ability, educational attainment, coronary artery disease, neuroticism, bipolar disorder type I, and anorexia nervosa.

Our identified GWS associations with fear were previously reported in GWASs of educational attainment [55], insomnia [56], major depressive disorder [57] and broadly-defined depression [58], including the *SNX29* gene. The sorting nexin family of proteins is implicated in neuronal function, synaptic plasticity, learning, and memory [59]. The SNP-level 14q24.3 locus and *PAX6* gene associated with fear were reported in GWAS of any anxiety disorder GWAS [20] and worry-neuroticism [60]. One GWS fear association was on the same locus (14q24.3) as the *YLPM1* gene associated with GAD. No other GWS fear associations overlapped with GAD results. As GWASs of specific anxiety disorders improve in power, we expect this overlap to increase. All GAD associations were identified in previous GWASs of distress-related phenotypes [19,20,57,58,60–63]. This includes the *SORCS3* gene, highly expressed in the CA1 hippocampus region and implicated in aversive memory extinction in mice [64], and the gut-brain axis [65]. Our finding supports further animal modelling and functional genomic research on *SORCS3* in distress liability.

High genetic correlations were observed between fear, GAD, and depression which reflects twin study findings [10,12,13]. Our results are compatible with our preliminary GWASs in the UK Biobank [22] but add detail to the genetic structure of fear and GAD by examining their relationship with a wide range of other complex traits. Screening controls specifically for the disorder/grouping being analysed improved our ability to differentiate genetic specificity between highly genetically correlated disorders. Evidence of some genetic specificity among fear and GAD emerged after screening controls specifically and excluding the depression-ascertained QIMR dataset. The inclusion of depression-enriched cases and controls screened broadly for other internalising disorders likely elevated shared internalising genetic liability, masking subtle genetic distinctions between fear and GAD. This highlights the importance of careful case-control ascertainment in identifying subtype-specific genetics.

Genetic correlations with depression were high and not significantly different between fear and GAD, supporting a common genetic liability factor shared among these disorders [10,12,13,66]. This common internalising liability factor is often conceptualised as being driven by shared negative affectivity, captured by measures of neuroticism [5,67]. We found neuroticism had a significantly stronger genetic correlation with GAD than fear. This is in line with twin studies showing genetic influences on GAD (and depression) were more core to a higher-order dimension of genetic liability to negative affectivity than fear [68]. General negative affectivity also influences fear-based disorders but is less central to them as they are characterised by more specific elements of acute fearful and physiological hyperarousal [4]. Our findings partially support a distress-fear genetic distinction but provide stronger evidence for an overarching internalising genetic liability driving comorbidity across anxiety disorders and depression.

Compared with GAD, fear had stronger negative genetic correlations with educational attainment and general cognitive ability, along with a higher positive genetic correlation with coronary artery disease (heart disease). The distinct genetic relationship between fear and cognitive-related traits may reflect shared cognitive mechanisms less central to GAD, as associative learning distinguishes fear-based disorders from GAD [69]. The unique genetic correlation of heart disease with fear aligns with a phenotypic study showing that fear-based disorders were stronger predictors of heart disease development than distress disorders [70]. This could be due to earlier onset and longer-term exposure to heart disease risk mechanisms in fear-based disorders [70–72]. Overlapping genetics with heart disease may underpin shared physiological pathways related to inflammation and autonomic dysfunction [73]. Low cardiac vagal tone may be a physiological mediator connecting anxiety and heart disease through maladaptive stress responses but is more consistently linked with panic disorder than GAD [74,75]. Over-self-reporting of heart disease (e.g., angina) [76] in fear-based disorders may partly explain our findings due to symptom overlap (e.g., chest pain) [70].

Our finding that GAD exhibited stronger positive genetic correlations than fear with bipolar disorder type I (BD-I), but not type II (BD-II), offers new insights into the anxiety-mood disorder genetic spectrum. While fear and GAD comorbidity rates are similar between BD-I and BD-II [77], our findings align with mood disorder genomics research. BD-II involves hypomania and is more similar to depression, whereas BD-I is characterised by severe manic episodes [78]. The mood disorder spectrum reflects a genetic continuum from manic to depressive clusters [79]. BD-I had a weaker genetic correlation with depression than BD-II and was more genetically similar to schizophrenia [80,81]. BD-I sits on one end of the genetic spectrum and recurrent major depressive disorder on the other, with BD-II closer to depressive clusters than BD-I, connecting the two [79]. Our novel finding adds detail to this genetic spectrum by highlighting heterogeneity at the internalising pole. Fear-based disorders lie at the opposite end to BD-I, with distress disorders (GAD and depression) sitting between bipolar and fear.

The weaker genetic correlation between fear and anorexia nervosa (∼0.07) compared to GAD (∼0.28) was unexpected given similar comorbidity rates, thought to be driven by shared genetic susceptibility and fear-conditioning mechanisms [82–84]. Further research is needed to establish whether genetic correlations with anorexia nervosa differ between individual fear-based disorders, with previous research restricted to analyses with GAD [85], or other eating disorder phenotypes [86,87]. A twin study suggests some individual fear disorder genetic specificity with broadly defined eating disorder, with a moderate genetic correlation with panic disorder, and lower genetic correlations with phobias and social anxiety disorder [87]. Shared loci affecting traits in different directions may hinder our ability to detect a *global* genetic correlation. *Local* genetic correlation analyses may identify specific shared genomic regions supporting common mechanisms linking fear-based disorders with anorexia nervosa [88,89].

Our findings should be considered in light of limitations. First, we were underpowered to assess individual fear-based disorders, which may have varying disorder-specific genetic influences as indicated by twin studies [12,13]. Second, combining panic disorder diagnosis with panic attack measures might have captured broad psychopathology genetics, given that panic attacks are transdiagnostic [78]. Third, specifically screening controls for a certain anxiety disorder was not possible in the QIMR due to limited measures. Fourth, combining detailed with brief measures may limit genetic specificity [28] and elevate the genetic correlation between fear, GAD, and related disorders like depression. Finally, we could not account for the high comorbidity observed across anxiety disorders and depression. In the GLAD+ dataset, only 5% of cases met criteria for a single anxiety disorder without comorbid MDD [3]. Ascertaining data for a well-powered GWAS of a single anxiety disorder without comorbidities will be challenging, given the considerable clinical overlap between anxiety disorders, and less representative of the broader lived experience of people with these disorders. Future studies with well-powered GWASs of anxiety subtypes with detailed phenotyping could discern subtype-specific genetics through multivariate genomic structural equation modelling [90]. As individual-level methods advance [91,92], this will enable the testing of genetic covariances across comorbid disorders and comparison of measures in smaller samples than is required for GWAS summary-level modelling.

In summary, our results provide further evidence for shared genetic liability between fear-based disorders, GAD, and depression, while revealing some genetic specificity. We identified differences in fear-GAD genetic relationships with general cognitive ability, coronary artery disease, neuroticism, bipolar type I and anorexia nervosa. Our findings add more fine-grained detail to the proposed hierarchical structure of internalising disorders and partially support fear-based disorders and GAD as separate distress-fear subtypes.

Quantitatively based dimensional modelling of shared and distinct distress-fear symptoms on the genomic level would further complete this hierarchical structure [5]. The growth of datasets with detailed phenotyping of all anxiety disorders, such as those used in this study, will be key to identifying subtype-specific and transdiagnostic genetic factors of the full anxiety disorder spectrum.

## Acknowledgements

We thank National Institute for Health and Care Research (NIHR) BioResource volunteers for their participation, and gratefully acknowledge NIHR BioResource centres, NHS Trusts and staff for their contribution. We thank the National Institute for Health and Care Research, NHS Blood and Transplant, and Health Data Research UK as part of the Digital Innovation Hub Programme. The views expressed are those of the author(s) and not necessarily those of the NHS, the NIHR or the Department of Health and Social Care. We gratefully acknowledge the participation of the NIHR BioResource Centre Maudsley, Biomedical Research Centre at South London and Maudsley NHS Foundation Trust and King’s College London volunteers and thank the staff for their help with volunteer recruitment. We thank the NIHR Biomedical Research Centre at South London and the Maudsley NHS Foundation Trust and King’s College London for funding. This study represents independent research supported by the NIHR Biomedical Research Centre BioResource at South London and Maudsley NHS Foundation Trust and King’s College London. We gratefully acknowledge capital equipment funding from the Maudsley Charity (Grant Ref. 980) and Guy’s and St Thomas’s Charity (Grant Ref. STR130505).

## Ethical approval

The London Fulham Research Ethics Committee approved the GLAD Study (21st August 2018; REC reference: 18/LO/1218) and EDGI UK (29th July, 2019; REC reference: 19/LO/1254). Ethical approval for the COPING Study was granted following a full review by the South West Central Bristol Research Ethics Committee (REC reference: 20/SW/0078) on 27th April 2020. The AGDS and QSkin cohorts were reviewed and approved by the QIMR Berghofer Medical Research Institute’s Human Research Ethics Committee. Analyses in the UK Biobank were performed under UK Biobank Project Application 82087. The UK Biobank has ethical approval from the North West Multi-centre Research Ethics Committee (MREC) as a Research Tissue Bank (RTB) approval. All participants provided informed consent.

## Author contributions

A.R.T.K, T.C.E and G.B. were responsible for the conception and design of the study. T.C.E, G.B, G.K., M.R.D, M.H., J.R.I.C and N.R.W, S.E.M, E.M.B, N.G.M established and provided access to the GLAD+ and QIMR datasets, respectively. A.R.T.K, C.H., A.J.P, H.L.D, J.M., Y.L., J.Z, A.P. were responsible for GLAD+ phenotypic data preparation and cleaning. A.R.T.K, S.H.L, J.R.I.C and B.A conducted preparation and quality control of genetic data. B.M carried out data analysis in the QIMR dataset. A.R.T.K conducted all other data analyses with support from G.M.V, S.H.L, H.L.D, J.M, M.S, J.R.I.C and A.E.F. A.R.T.K and T.C.E were responsible for the drafting of the original manuscript. All co-authors reviewed the manuscript.

## Data availability

Fear and GAD GWAS summary statistics will be made publicly available upon publication on Zenodo. Access to the individual-level data used in this study is subject to approval by the respective data access committees and can be requested as follows: GLAD, COPING and EDGI UK through the NIHR BioResource at https://www.bioresource.nihr.ac.uk/using-our-bioresource/apply-for-bioresource-data-access; UK Biobank at https://www.ukbiobank.ac.uk/enable-your-research/apply-for-access/; AGDS data access requests should be directed to N.G.M.

## Code availability

The analyses were conducted using the following publicly available software: REGENIE (v3.1.3) (https://github.com/rgcgithub/regenie), SAIGE (v0.44) (https://github.com/weizhouUMICH/SAIGE), METAL (2020-05-05 release) (https://github.com/statgen/METAL), MungeSumstats (v1.10.1) (https://github.com/Al-Murphy/MungeSumstats), FUMA (v1.5.2) (https://fuma.ctglab.nl/) and LDSC (v1.0.1) (https://github.com/bulik/ldsc).

## Consortia

**NIHR BioResource group author**: John Bradley, Nathalie Kingston, Hannah Stark, Carola Kanz, Alexei Moulton, Nigel Ovington, Jacinta Lee, Debbie Clapham-Riley, Katie Mills.

